# Outcomes of open transverse abdominis release for ventral hernias- A systematic review, meta-analysis and meta-regression of factors affecting them

**DOI:** 10.1101/2022.05.12.22275032

**Authors:** Bhavin B Vasavada, Hardik Patel

## Abstract

**Objectives:** The primary objectives were to evaluate Surgical Site Occurrences (SSO) and Surgical Site Occurrences requiring procedural Intervention (SSOPI) after open transversus abdominis release and to study various factors affecting it. Secondary objectives were to evaluate Surgical Site Infections (SSI), recurrence rates and overall complications after transversus abdominis release (TAR) and the factors responsible for those.

**Methods:** We searched PUBMED, SCOPUS and Cochrane databases with keywords “transversus abdominis release” or “TAR” OR “Surgical Site Occurrences” OR “posterior component separation AND “outcomes” as per PRISMA 2020 and MOOSE guidelines. Full texts and English literature studies were included, studies mentioning outcomes for open transversus abdominis release for ventral hernia were included and studies with robotic transversus abdominis release were excluded. Percentage occurrences of SSO, SSOPI, SSI, recurrence and overall complications after TAR were evaluated. Random effect meta-analysis with restricted maximum likehood methods was used for meta-analysis. Heterogeneity was analysed using I^2^ statistics. Publication bias with eager’s test and funnel plots. Meta0regression analysis was done to evaluate factors affecting the heterogeneity. JASP 0.16.2 software was used for meta-analysis.

**Results:** Twenty two studies including 5284 patients who underwent TAR for ventral hernia were included in systematic review and meta-analysis. Overall pooled SSO, SSOPI, Overall Complications, SSI and recurrence rates were 21.72% [95% C.I 17.18-26.27%], 9.82% [95% C.I 7.64 −12%], 33.34% [95% C.I. 27.43-39.26%], 9.13% [95% 6.41-11.84] and 1.6% [0.78-2.44] respectively. Heterogeneity was significant in all the analysis. Age (p<0.001),sex (p<0.001), BMI (p<0.001),presence of comorbidities (p<0.001), prior recurrence, defect size (p<0.001) and current or past history of tobacco exposure were associated with SSO in multivariate meta-regression analysis. Defect size (p=0.04) was associated with SSOPI. Age (p=0.011), BMI (p=0.013), comorbidities (p<0.01), tobacco exposure (p=0.018),prior recurrence (p <0.01) and sex (p < 0.01) were associated with overall complications.

**Conclusion:** Open transversus abdominis release is associated with high rates of SSO, SSOPI, SSI and overall complications but recurrence rates are low. Various preoperative factors mentioned may be responsible for heterogeneity across studies.

## Background

Ventral and Incisional hernias are one of the most complex hernias with high rates of complications as well as recurrences. To deal with complex ventral hernias component separation technique was first described by Ramirez et al. [1]. After that many techniques have been described to deal with this complex problem.

Transversus abdominis release is a recent procedure that has shown excellent results in repairing the complex ventral or incisional hernia. However, it is still associated with high complication rates. [2]. Very few studies review various complications after transversus abdominis release and much lesser studies in the literature evaluate factors responsible for various complications.

There are certain standard terms to describe commonest complications like wound complications after ventral or incisional hernia repair. Ventral Hernia Working Group 2010 coined the term Surgical Site Occurrences. (SSO) [3] The term Surgical Site Occurrences requiring Procedural Interventions (SSOPI) has been introduced recently. [4]

The primary objectives of this systematic review and meta-analysis were to evaluate Surgical Site Occurrences (SSO) and Surgical Site Occurrences requiring procedural Intervention (SSOPI) after open transversus abdominis release and to study various factors affecting it. Secondary objectives were to evaluate Surgical Site Infections (SSI), recurrence rates and overall complications after transversus abdominis release (TAR) and the factors responsible for those.

## Material and Methods

The study was conducted according to the Preferred Reporting Items for Systematic Reviews and Meta-Analyses (PRISMA) 2020 statement and MOOSE guidelines. [5,6]. We conducted a literature search as described by Gossen et al. [7]. We searched PubMed, Scopus, and Cochrane databases with keywords “transversus abdominis release” or “TAR” OR “Surgical Site Occurrences” OR “posterior component separation AND “outcomes”. Two independent authors extracted the data (B.V and H.P). In case of disagreements, a decision is reached on basis of discussion.

### Definitions

We defined surgical site occurrences, surgical site occurrences requiring procedural interventions and surgical site infections as per DeBoard et al. [8].

Surgical Site Occurrences: SSI, seroma, wound dehiscence, enterocutaneous fistula, wound cellulitis, non-healing incisional wound, fascial disruption, skin or soft tissue ischemia, skin or soft tissue necrosis, wound serous or purulent drainage, stitch abscess, seroma, hematoma, and infected or exposed mesh.

Surgical Site Occurrences Requiring Procedural Intervention: SSOs require a procedural intervention, defined as wound opening or debridement, suture excision, percutaneous drainage, or mesh removal.

Surgical Site Infection: Infection occurring where the surgery took place and includes superficial deep, and organ space infections

Overall complications were defined as all wound complications mentioned above plus all the other systematic complications mentioned in the studies included.

Recurrence was defined as any recurrences within 30 days mentioned in included studies.

Preoperative horizontal defect size was taken as defect size.

Inclusion criteria for studies.

1. Studies evaluating open transversus abdominis release outcomes

2. English language studies

3. Full texts.

4. Studies mention various preoperative and intraoperative variables.

### Exclusion criteria

1. Abstracts only

2. Articles other than the English language

3. Studies mention only minimal invasive transversus abdominis release.

4. Studies where full texts could not be obtained.

### Statistical Analysis

This meta-analysis was done with the JASP Team (2020). JASP (Version 0.14.2)(University of Amsterdam). Percentage rates of SSO, SSOPI, SSI, overall complications and 30 days recurrences were included as effect sizes to get weighted percentage rates as summary effects. Standard errors were calculated manually. A random-effect meta-analysis with Restricted Maximum Likehood methods was used. Multivariate meta-regression models were used to analyse which factors independently were associated with heterogeneity and indirectly to summary effects. Heterogeneity was assessed using the Higgins I^2^ test, with values of 25%, 50%, and 75% indicating low, moderate, and high degrees of heterogeneity, respectively. [9]. we also assessed the p-value for the significance of heterogeneity and tau^2^ and H^2^ values whenever possible. Publication bias was analysed using funnel plots and eager’s test.

### Summary of Bias

Cohort studies were assessed for bias using the Newcastle-Ottawa Scale to assess the risk of bias. [10].

## Results

“PUBMED”, “SCOPUS”, and “COCHRANE” databases were searched using the above keywords. Search strategy as per PRISMA statement 2020 is described in Figure 1. Twenty-two studies including 5248 patients were included in the study.[11–32] We have described study characteristics in Table 1. A summary of bias with the Newcastle-Ottawa scale is included in table 2.

**Table 1.**
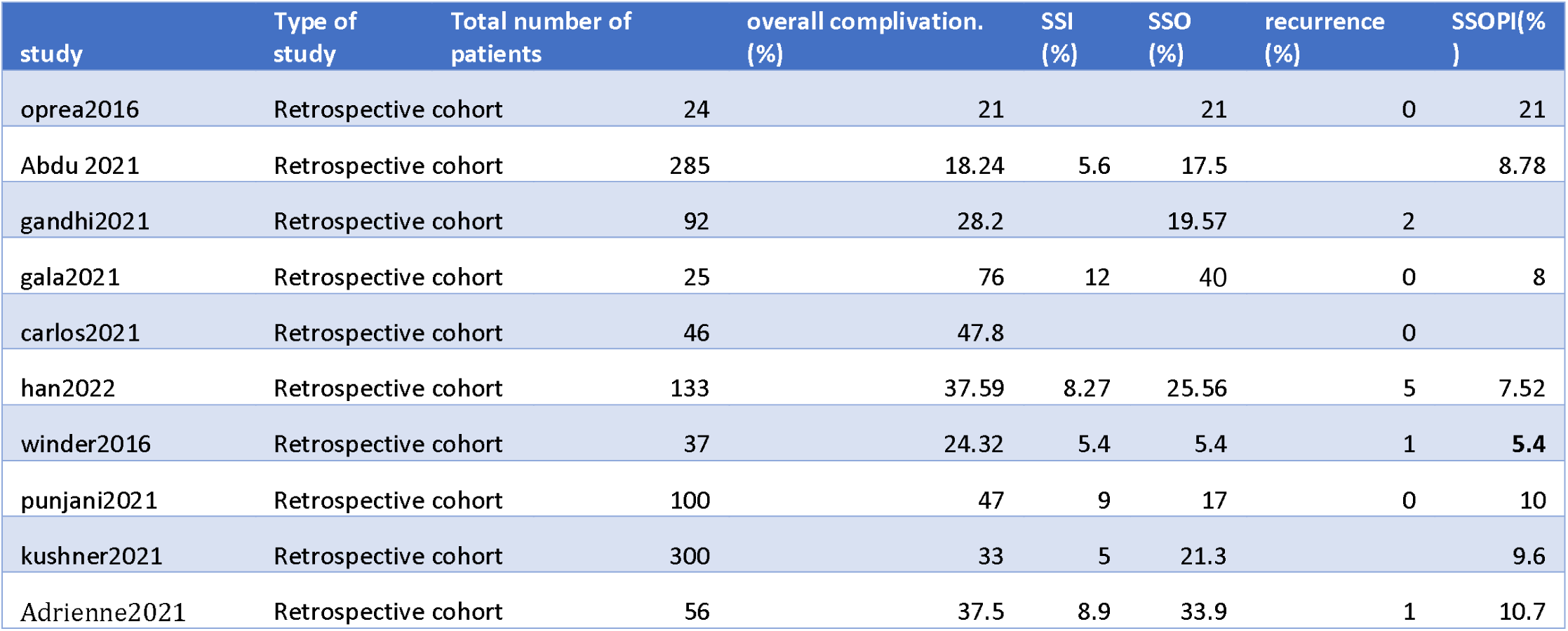
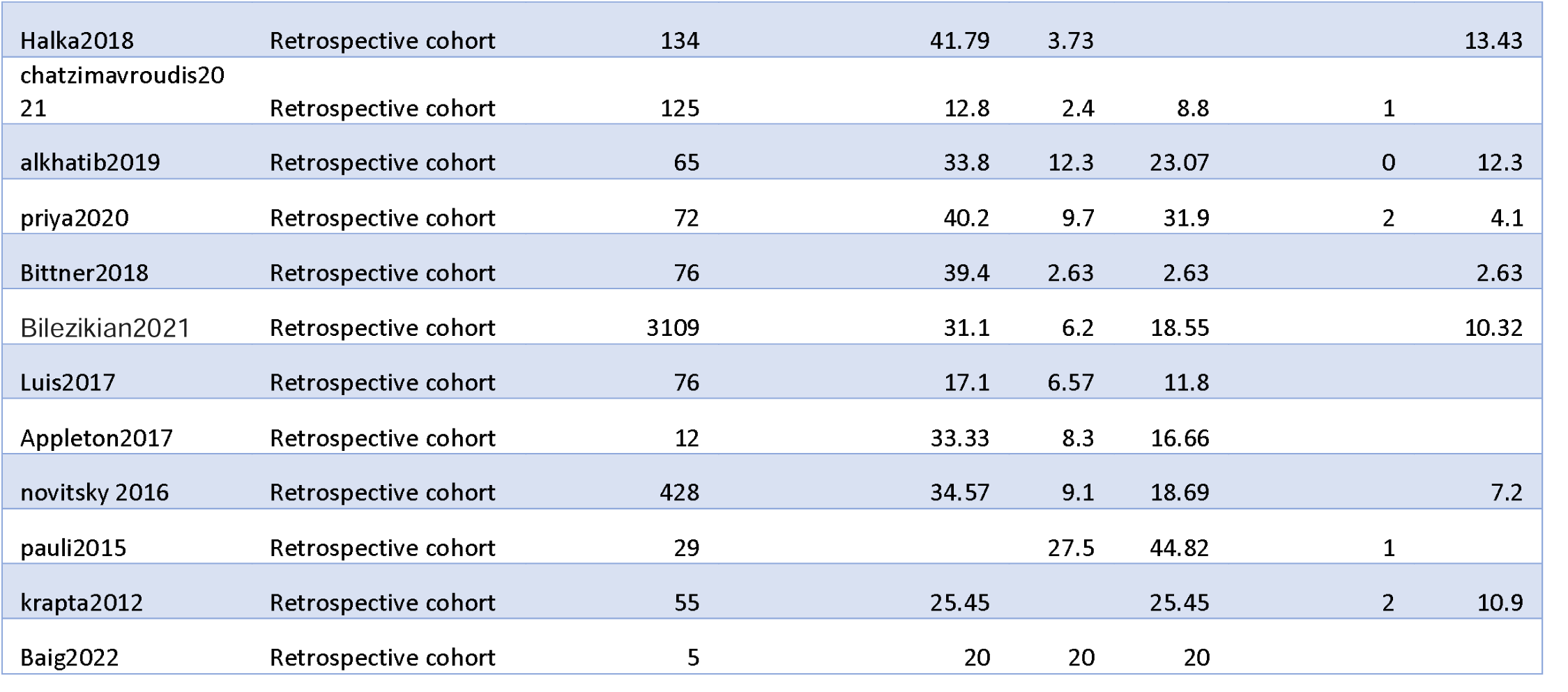
Study Characteristics.

**Table 2:** Summary of Bias. NewCastle- Ottawa scale.

| NOS for the risk of bias and quality assessment of Non Randomised Studies |  |  |  |  |  |  |  |  |  |  |
| --- | --- | --- | --- | --- | --- | --- | --- | --- | --- | --- |
| Author | Year | Selection |  |  |  | Compa<br>rability | outcomes |  |  | To<br>tal<br>sc<br>or<br>e |
|  |  | Represe<br>ntative<br>of<br>expose<br>d<br>cohort | sele<br>ction<br>of<br>non<br>ex<br>posed<br>coh<br>orts | Ascerta<br>inment<br>of<br>exposu<br>re | Demon<br>stration<br>of<br>outcom<br>e of<br>interest<br>not<br>present<br>at start | compar<br>ibility<br>of<br>cohort<br>based<br>on<br>design<br>and<br>analysi<br>s | Asses<br>ment<br>of<br>outco<br>mes | was<br>foll<br>ow<br>up<br>eno<br>ugh<br>for<br>outc<br>ome<br>to<br>occ<br>ur? | adeq<br>uacy<br>of<br>foll<br>ow<br>up<br>of<br>coho<br>rts |  |
| Opera.<br>[11] | 2016 | ★ |  | * | ★ | ★ | ★ | ★ | * | 7 |
| Abdu et<br>al. [12] | 2021 | ★ |  | * | * | * | ★ | ★ |  | 6 |
| gandhi et<br>al. [13] | 2021 | ★ |  | * | ★ | ★ | ★ | ★ | * | 7 |
| Gala et<br>al. [14] | 2021 | ★ |  | * | ★ | ★ |  | ★ | * | 6 |
| carlos et<br>al. [15] | 2021 | ★ |  | * | ★ | ★ | ★ | ★ | * | 7 |
| Han et al.<br>[16] | 2022 | ★ |  | * | ★ | ★ | ★ | ★ | * | 7 |
| Winder<br>et al. [17] | 2016 | * |  | * | * | * | * | * | * | 7 |
| Punjani | 2021 | * |  | * | * | * | * | * | * | 7 |
| et al. [18] |  |  |  |  |  |  |  |  |  |  |
| Kushner<br>et al. [19] | 2021 | * |  | * | * | * | * | * |  | 6 |
| Adrienne<br>et al. [20] | 2021 | * |  | * | * | * | * | * | * | 7 |
| Halka et<br>al. [21] | 2018 | * |  | * | * | * |  | * |  | 5 |
| chatzima<br>vroudis<br>et al. [22] | 2021 | * |  | * | * | * |  | * | * | 6 |
| Alkhatib<br>et al. [23] | 2019 | * |  | * | * | * | * | * | * | 7 |
| Priya et<br>al. [24] | 2020 | * |  | * | * |  |  | * | * | 5 |
| Bittner et<br>al.[25] | 2018 | * |  | * | * | * | * | * |  | 6 |
| Bilezikia<br>n et al.<br>[26] | 2021 | * |  | * | * | * | * | * |  | 6 |
| Luis et<br>al. [27] | 2018 | * |  | * | * | * |  | * |  | 5 |
| Appleton<br>et al. [28] | 2017 | * |  | * | * | * |  | * |  | 5 |
| Novitsky<br>et al. [29] | 2016 | * |  | * | * | * | * | * |  | 6 |
| Pauli et<br>al. [30] | 2015 | * |  | * | * | * |  | * |  | 5 |
| Krapta et<br>al. [31] | 2012 | * |  | * | * | * | * | * | * | 7 |
| Baig et<br>al. [32] | 2022 | * |  | * | * |  |  | * |  | 4 |
| NOS, Newcastle-Ottawa scale.. |  |  |  |  |  |  |  |  |  |  |

### Surgical Site Occurrences (SSO)

Overall weighted surgical site occurrences rate was 21.72% with 95% C. I 17.18-26.27%. Heterogeneity was high with I^2^ 100% and significant. (p<0.001). Publication bias was nonsignificant with Egger’s test. (p= 0.358). The Forest plot for Surgical Site Occurrences and Funnel plot is included in Figure 2. Age (p<0.001),sex (p<0.001), BMI (p<0.001),presence of comorbidities (p<0.001), prior recurrence, defect size (p<0.001) and current or past history of tobacco exposure (p <0.001) were independently associated heterogeneity and so indirectly with SSO in multivariate meta-regression analysis.

### Surgical Site Occurrences requiring procedural intervention: (SSOPI)

Over weighted SSOPI rate was 9.82% with a 95% confidence interval of 7.64-11.99%. Heterogeneity was high with I^2^ 100% and significant. (p<0.001). Publication bias was significant with Egger’s test. (p=0.04). Forest plots for SSOPI and Funnel plot for publication bias are shown in figure 3. On multivariate metaregression analysis, preoperative defect size independently predicted heterogeneity and indirectly SSOPI. (p=0.04).

### Overall Complications

The weighted overall postoperative complication rate was 33.34% with a 95% confidence interval of 27.43-39.26%. Heterogeneity was high with I^2^ 100% and statistically significant. (p <0.001). Egger’s test for publication bias was nonsignificant. (p=0.831). Age (p=0.011), BMI (p=0.013), comorbidities (p<0.01), tobacco exposure (p=0.018),prior recurrence (p <0.01) and sex (p < 0.01) were independently associated with heterogeneity and indirectly with overall complications. The Forest Plot for a summary of effect and the Funnel plot for publication bias is shown in figure 4.

### Surgical Site Infection. (SSI)

The weighted Overall Surgical Site Infection rate was 9.13% with 95% Confidence intervals of 6.41%-11.84%. Heterogeneity was high and significant with I^2^ 99.99% and statistically significant. (p<0.001). Egger’s test for publication bias was nonsignificant. (p=0.127). The Forest Plot for a summary of effect and the Funnel plot for publication bias is shown in figure 5. No Factors were independently associated with Heterogeneity in the multivariate meta-analysis.

### Hernia recurrence

Weighted overall recurrence rate was 1.61% with 95% confidence interval 0.78- 2.44%. Heterogeneity was high with I^2^ 99.99% and statistically significant. (p<0.001). Egger’s test for publication bias was statistically significant. (p < 0.001). The Forest Plot for a summary of effect and the Funnel plot for publication bias is shown in figure 6. Preoperative defect size was associated with Heterogeneity and indirectly with hernia recurrence. (p=0.048).

## Discussion

One of the key goals of complex ventral or incision hernia repair is tension-free closure of midline fascia with mesh, which can reduce the recurrence rate. [33]. Ventral hernias with larger defects > 10 cm and chronic hernias are recently treated with the transverse abdominis release technique. However, this technique is associated with higher percentages of wounds and other complications. Various factors are associated with overall and wound complications[34].

This systematic review and meta-analysis aim to analyse wound-related complications like surgical site occurrences and surgical site occurrences requiring procedural interventions defined as above after open TAR. We also evaluated surgical site infections, recurrences and overall complications. We also did meta-regression analysis to study various factors associated with heterogeneity between studies and hence indirectly with the above complications.

The results of this meta-analysis show that various wound-related and overall complications remain high with SSO, SSOPI, and SSI and overall complication rates are around 21.72%,9.82%, 9.13% and 33.14% respectively but recurrences rates after TAR remain low with 1.61%. This confirms that TAR is a very effective procedure for complex ventral hernias but morbidity after TAR remains high and TAR should be performed after proper evaluation of indications.

On metaregression analysis age, sex, BMI, prior recurrence, associated comorbidities, defect size and current or past exposure to tobacco or smoking were independently associated with heterogeneity and indirectly with SSO. Most of the above factors were also associated with heterogeneity between studies for overall complications, which shows that patient-related factors were associated with SSO and overall complications. However, for SSOPI only defect size was independently associated with heterogeneity between the studies, which shows SSOPI may be associated with large and complex hernias. Defect size was also associated with heterogeneity in recurrence analysis. Which shows complex and larger ventral hernia requires postoperative interventions and also recurrences are common with them.

We analyzed open TAR, in studies which mentioned both open and robotic data [12,21,24,25,27], we included their data for open TAR in the analysis. The recent meta-analysis comparing outcomes of open vs robotic TAR [35], concluded that though overall complications and SSO rates are low in robotic TAR, the SSOPI, SSI, reoperation and readmission rates are similar. We need further studies regarding cost-benefit ratios comparing open vs robotic TAR.

To our knowledge this is the first meta-analysis with meta-regression, studying various factors responsible for heterogeneity in various outcomes in different studies, it is also the meta-analysis including the highest number of patients till now.

The limitations of this meta-analysis are some studies did not describe all the outcomes of interest, some did not include all the factors studied in the meta-analysis, and we could not include various factors like operative time, prior number of surgeries, prior mesh infection, prior contaminated or dirty wound as enough number of studies did not mention these factor to run meta-regression analysis. Heterogeneity was high and significant in most of the analyses, residual heterogeneity was also significant after meta-regression analysis. So, we cannot exclude some other confounding factors responsible for heterogeneity. However, publication bias was not significant in most of the analyses.

In conclusion, though open transversus abdominis release surgery is a highly effective procedure with low recurrence rates in complex ventral/incisional hernias, morbidity and wound complications remain higher. Various patient and hernia related factors are associated with SSO, SSOPI and overall complications. Proper selection of patients is the key to satisfactory outcomes after open transversus abdominis release surgery.

## Data Availability

All data produced in the present study are available upon reasonable request to the authors

## Abbreviations

SSO: Surgical Site Occurrences
SSOPI: Surgical Site Occurrences requiring Procedural Intervention
SSI: Surgical site infection
(TAR): Transversus abdominis release

---

**PRISMA 2020 flow diagram for new systematic reviews which included searches of databases and registers only**

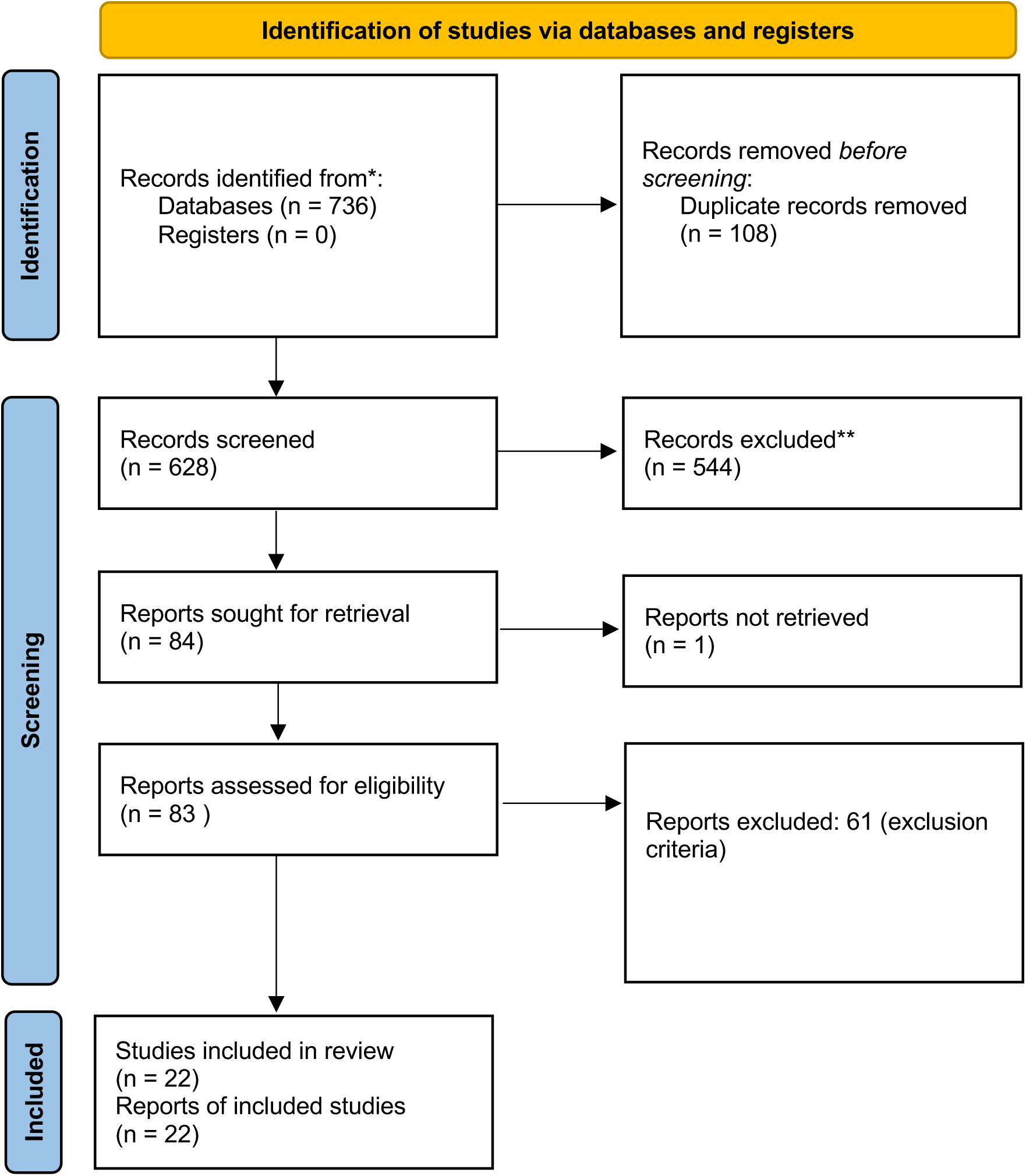

*From:* Page MJ, McKenzie JE, Bossuyt PM, Boutron I, Hoffmann TC, Mulrow CD, et al. The PRISMA 2020 statement: an updated guideline for reporting systematic reviews. BMJ 2021;372:n71. doi: 10.1136/bmj.n71

For more information, visit: http://www.prisma-statement.org/

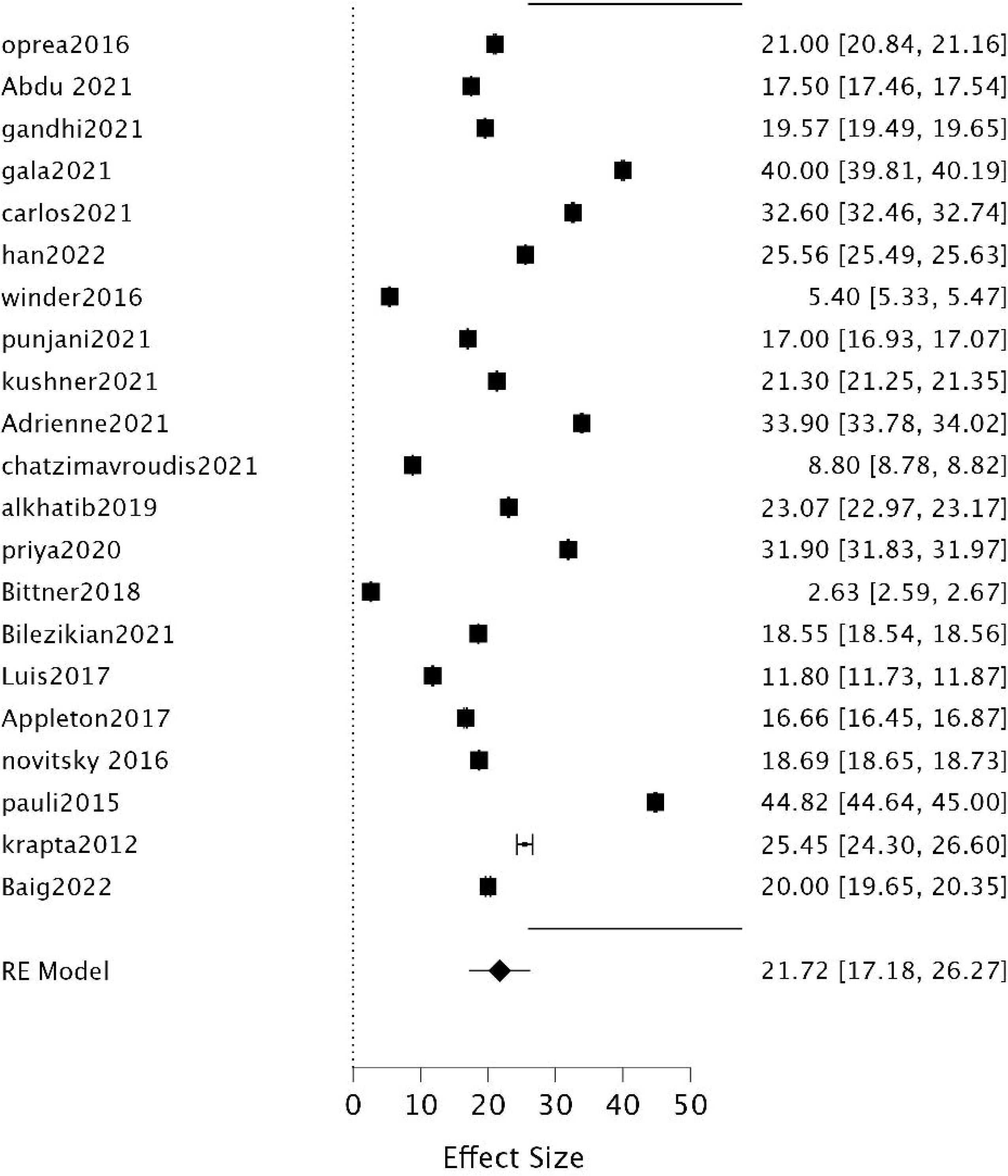

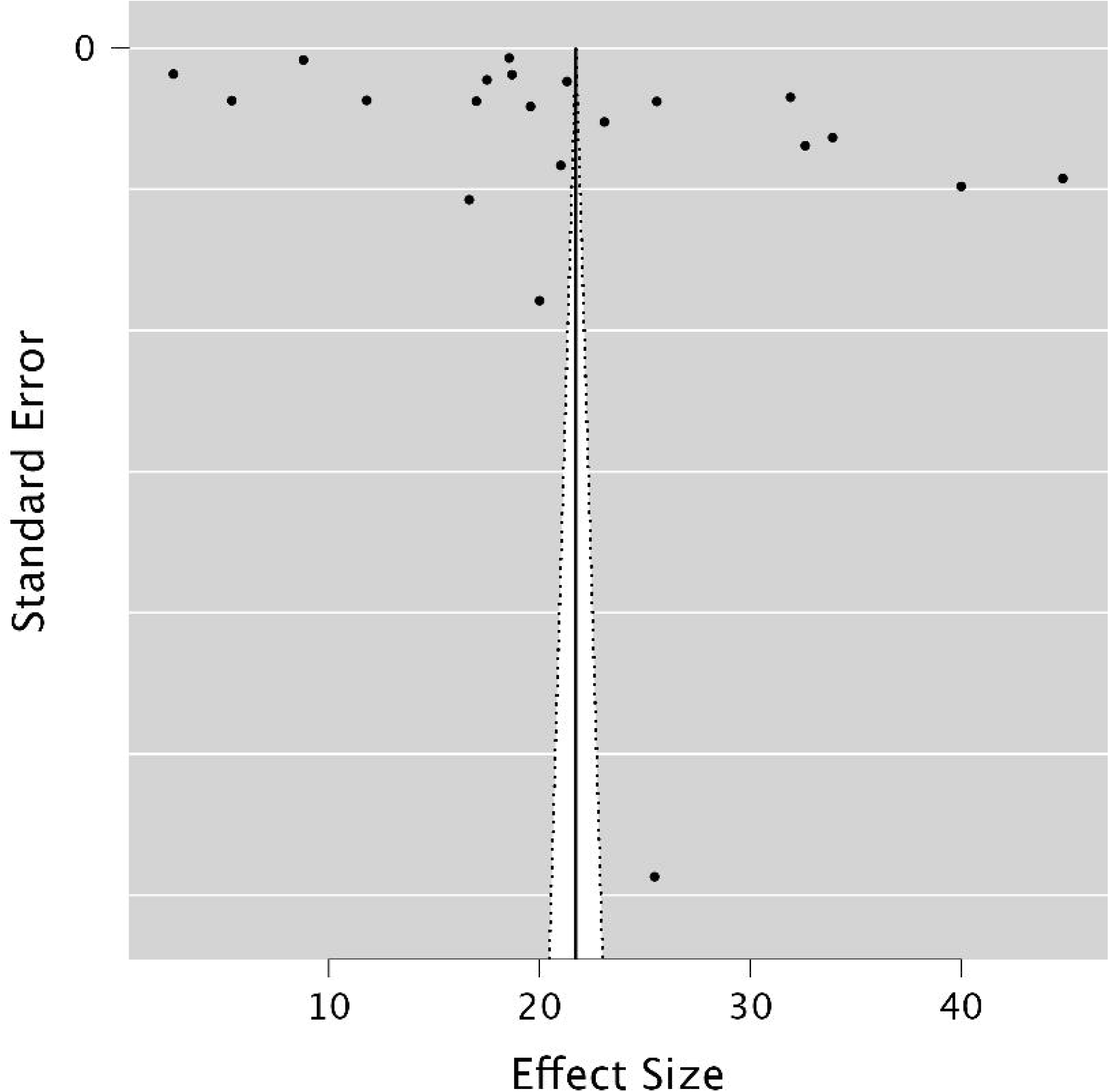

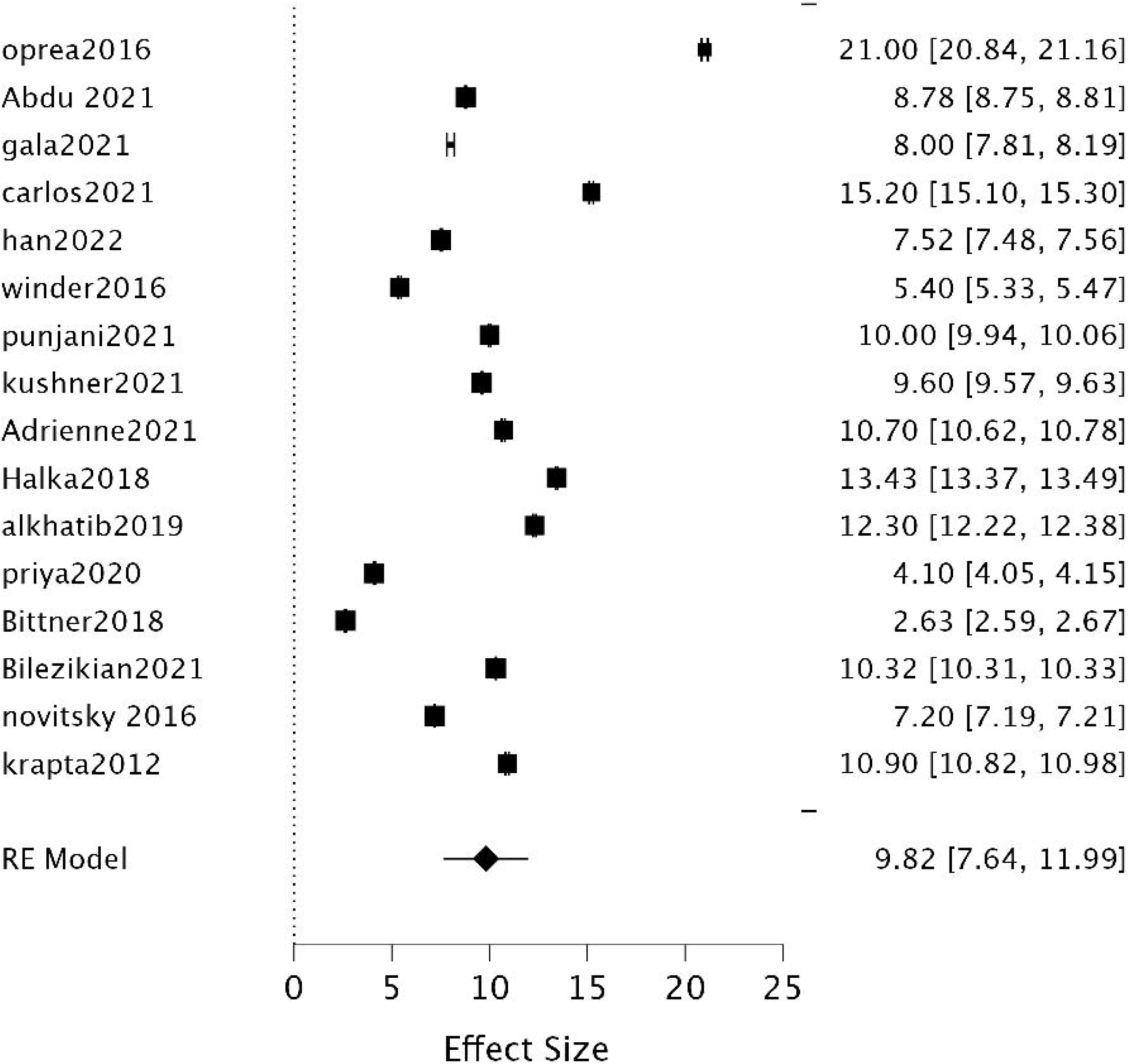

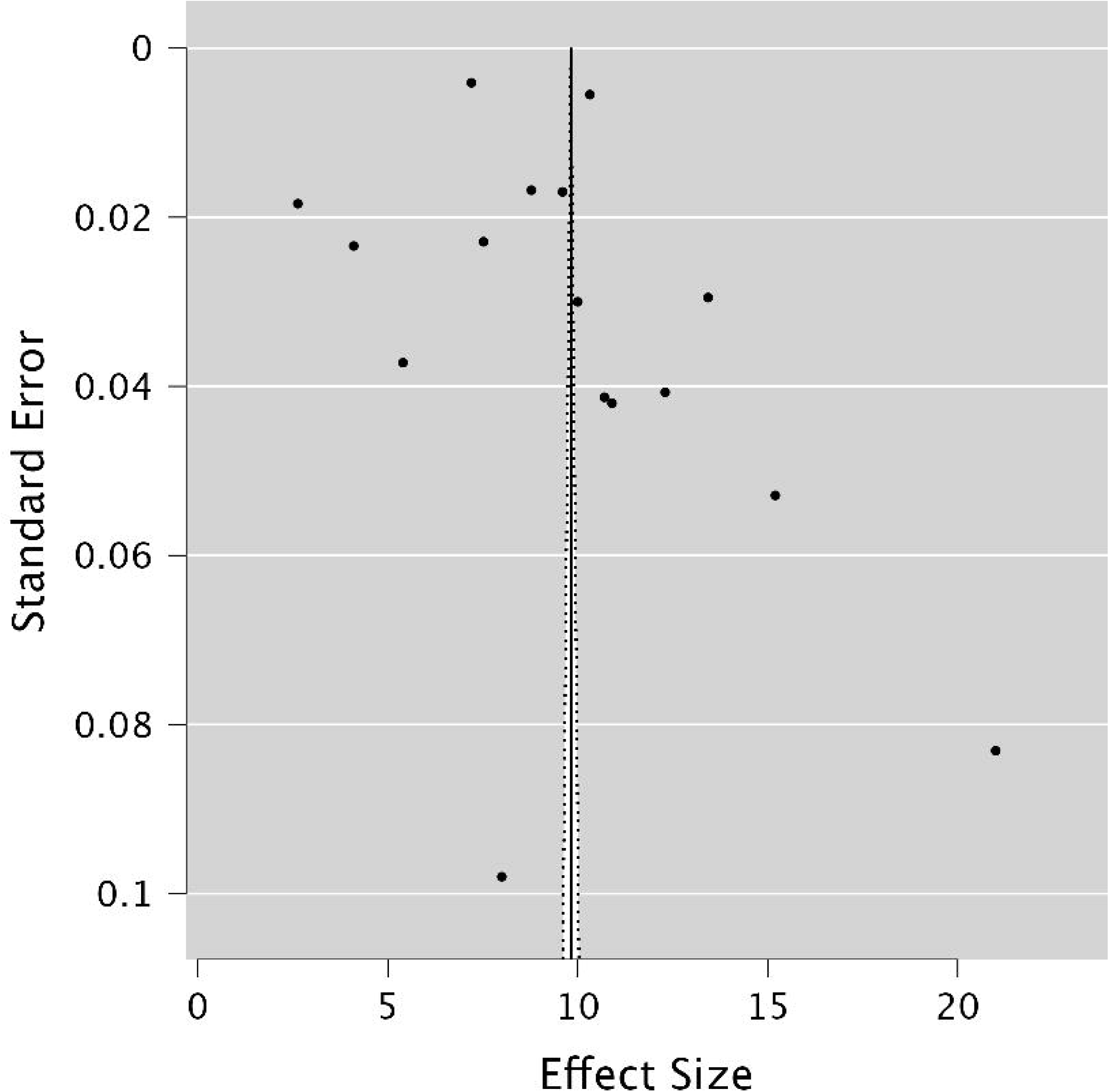

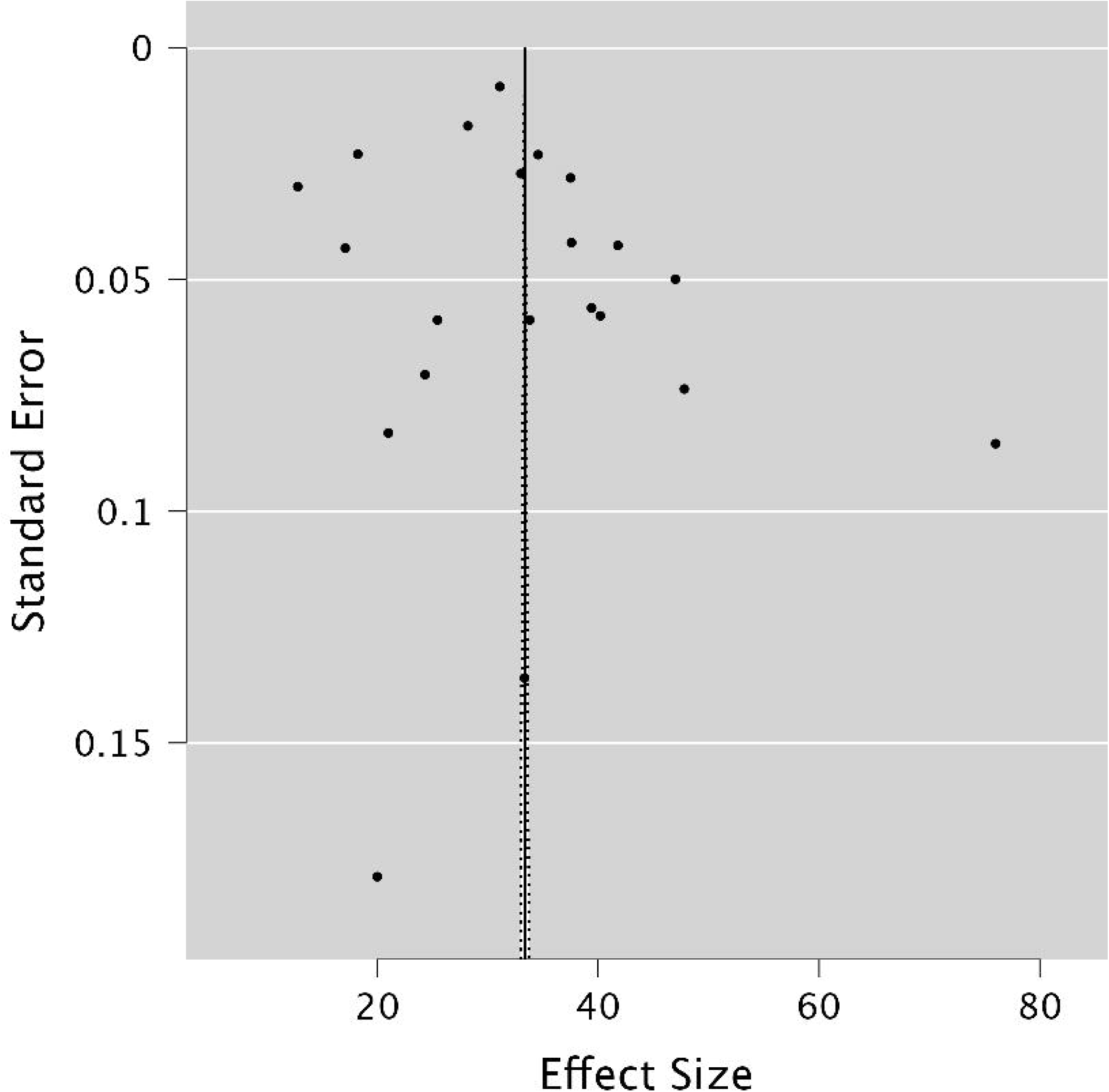

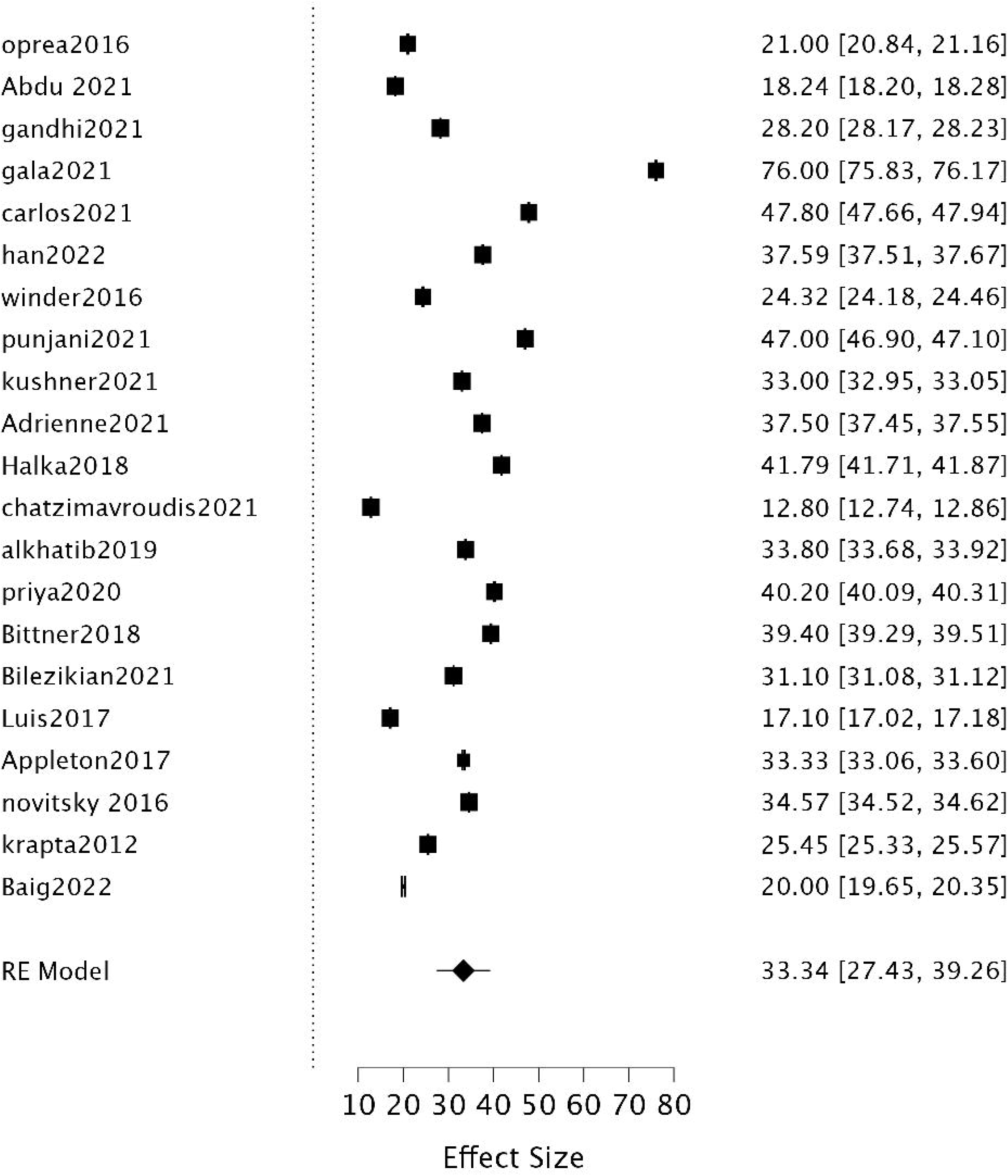

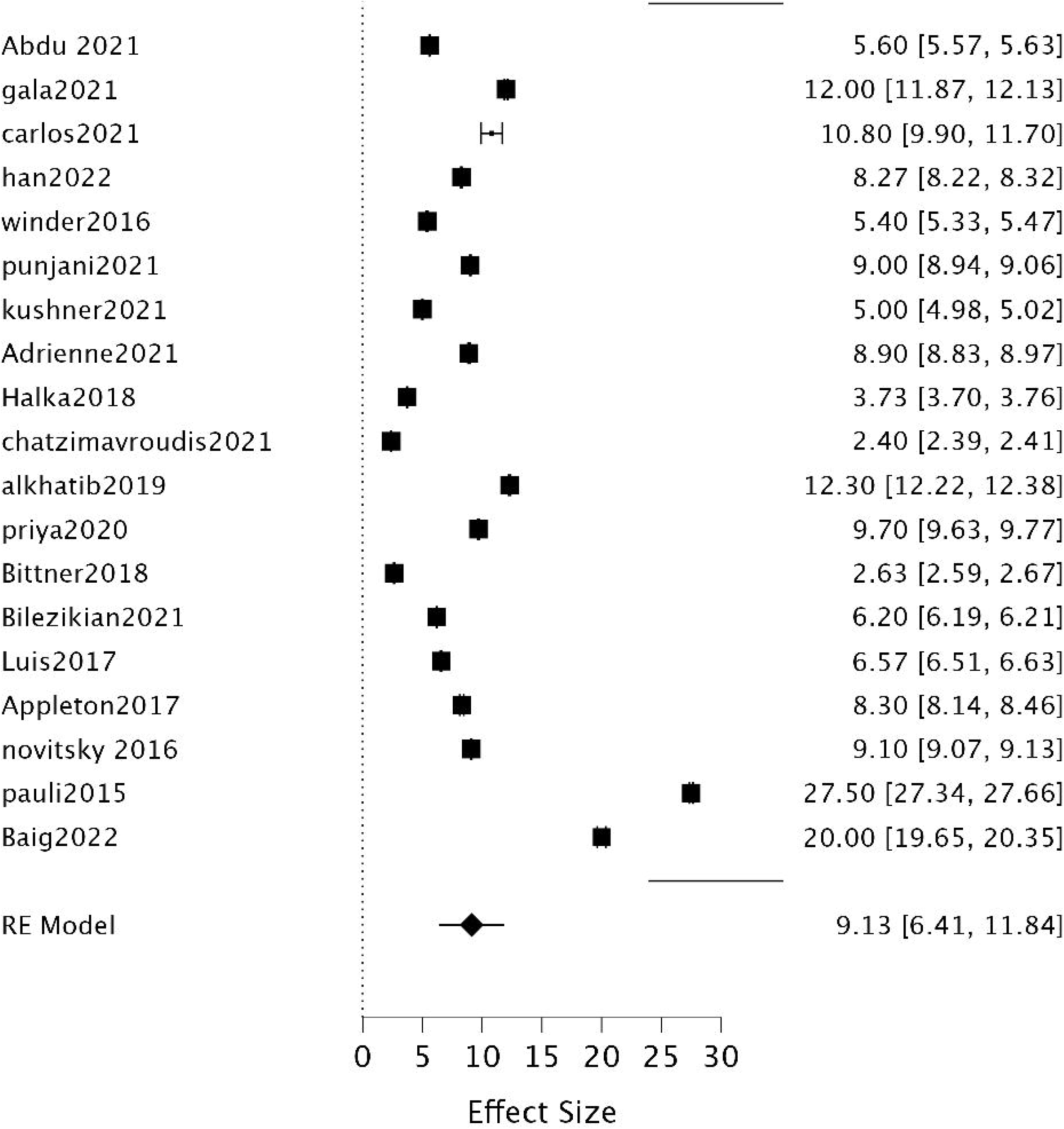

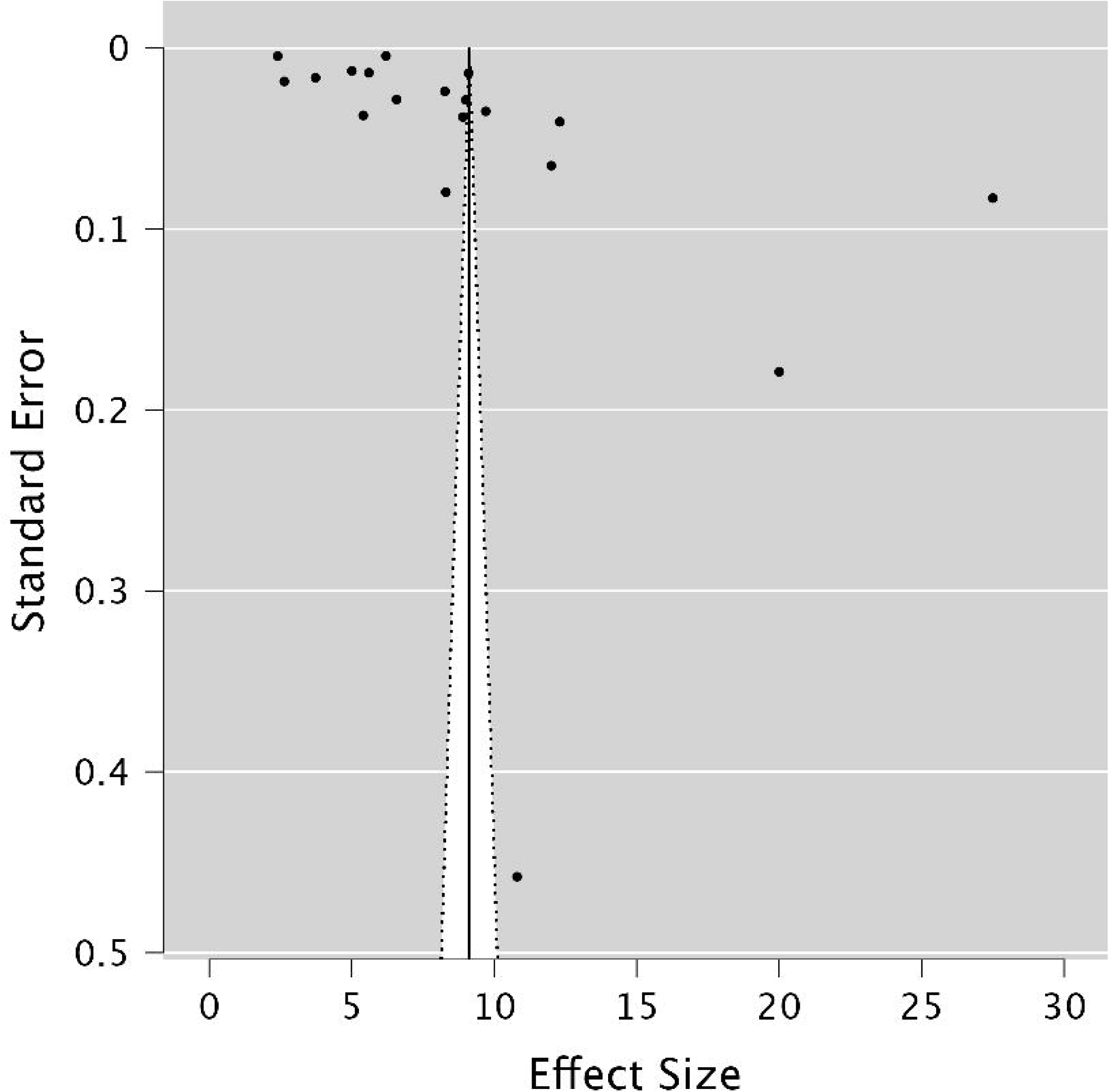

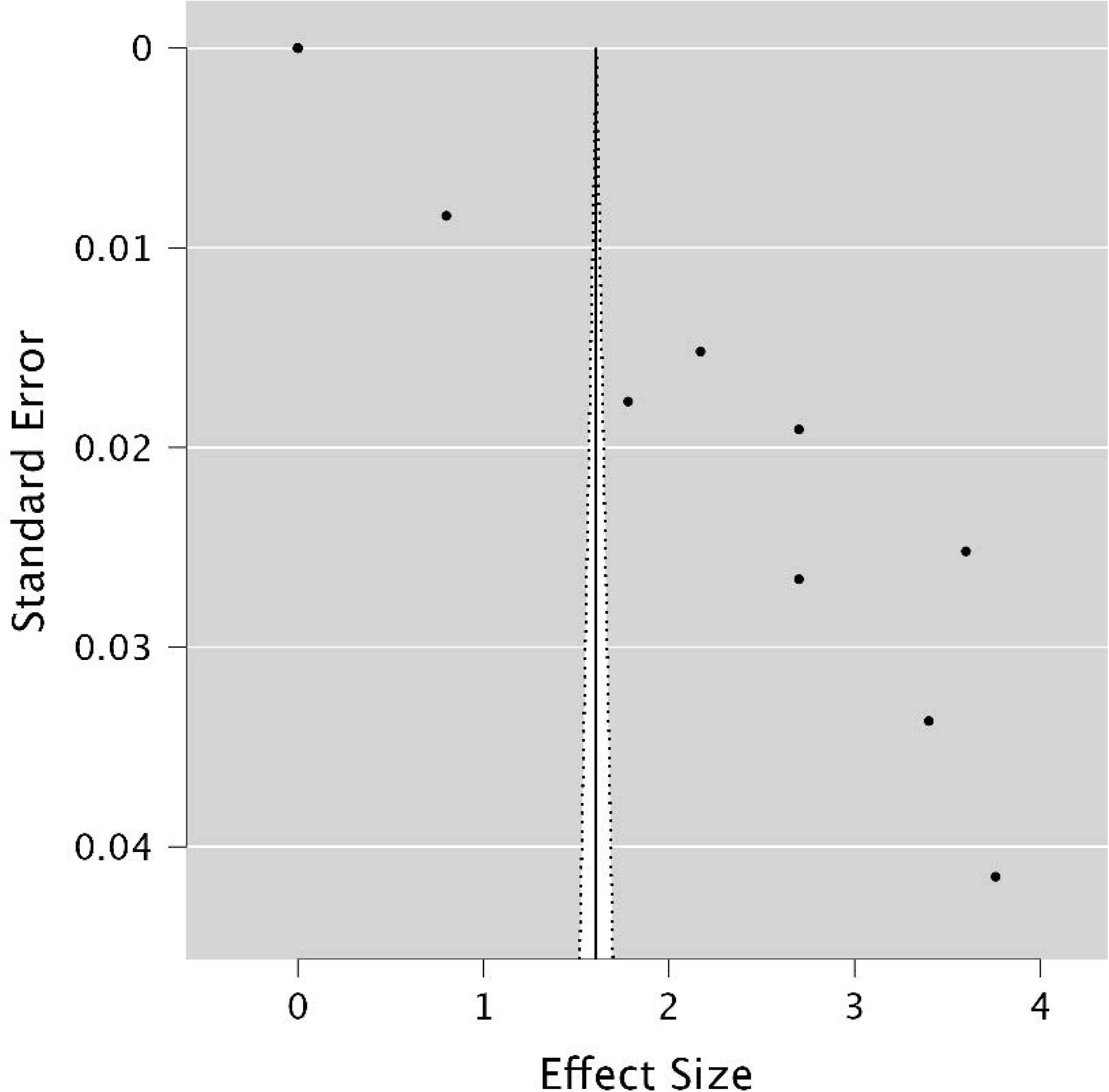

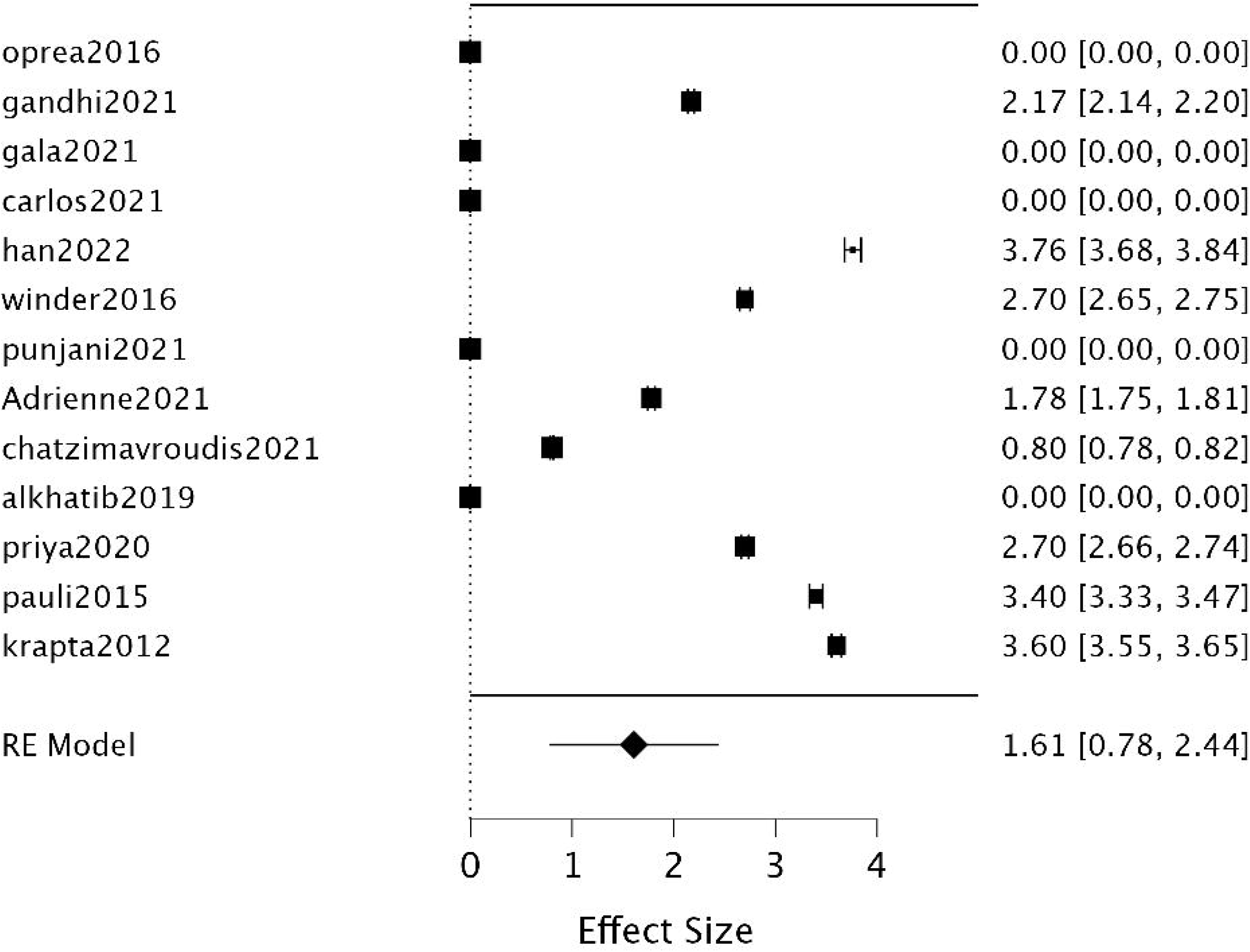

## Notes

Conflict of interests: none

### Competing Interest Statement

The authors have declared no competing interest.

### Funding Statement

This study did not receive any funding

